# Alzheimer’s neuropathology and longitudinal change in subjective cognitive complaints

**DOI:** 10.1101/2024.09.25.24314211

**Authors:** Alisa Bannerjee, Wendy Elkins, Susan M. Resnick, Murat Bilgel

## Abstract

**INTRODUCTION:** Subjective cognitive complaints often precede declines in objective measures of cognitive performance. Associations of incipient AD neuropathology with subjective cognitive complaints may be detectable earlier than its associations with neuropsychological testing among cognitively normal individuals.

**METHODS:** We examined the independent associations of PET measures of amyloid-β and tau pathologies with longitudinal subjective complaints and memory among 91 cognitively normal Baltimore Longitudinal Study of Aging participants using linear mixed effects models. Subjective complaints and memory performance were assessed with the Cognitive Failures Questionnaire and the California Verbal Learning Test, respectively.

**RESULTS:** Greater parahippocampal tau, independent of amyloid, was associated with higher subjective complaints (estimate=0.25, SE=0.1, *p*=0.015), while greater entorhinal tau corresponded to an attenuated increase in complaints over time (estimate=−0.06, SE=0.03, *p*=0.047). Hippocampal tau was associated with steeper memory decline (estimate=−0.03, SE=0.01, *p*=0.040).

**CONCLUSION:** Subjective cognitive complaints may be a reflection of early cerebral tau pathology in cognitively normal individuals.

## 1 Introduction

Subjective cognitive complaints are individuals’ perceived and self-reported memory complaints, or memory complaints reported by a close relative or friend who has observed changes in the individual’s cognition [1]. The 2018 National Institute of Aging – Alzheimer’s Association Alzheimer’s Disease (AD) Research Framework [2] as well as the Alzheimer’s Association Workgroup’s revised criteria for diagnosis and staging of Alzheimer’s disease [3] identify clinical stage 2 as the time when an individual is experiencing subtle cognitive change, which can be measured by subjective cognitive complaints, but does not show cognitive impairment on objective testing. Some studies have shown that participants reporting higher subjective cognitive complaints had worse scores on neuropsychological testing [4,5] and faster rates of decline in neuropsychological testing [6]. Participants with subjective cognitive complaints are at a higher risk for progressing to dementia versus those without these complaints [7–9]. However, other studies did not find an association between subjective cognitive complaints and objective cognitive assessments [10,11].

The revised criteria for AD suggest that for individuals in clinical stage 2, defined by subtle cognitive decline including subjective decline, the typical expected biological stage is B, which is characterized by the presence of amyloid-β plaques and medial temporal tau [3]. This is supported by research showing that individuals with subjective cognitive complaints have higher levels of amyloid and tau in the brain [8]. Detecting these neuropathological changes in early stages is essential as studies have shown that the onset of these changes precede the onset of clinical AD symptoms by up to 15 years for CSF tau and 20 years for CSF Aβ_42_ [12,13]. Furthermore, a study using data from ADNI found that the median time from estimated amyloid onset to clinical AD symptoms, as determined with a Clinical Dementia Rating (CDR) ≥ 1, to be approximately 14 years [14].

Prior research has offered insight into the association between neuropathology and subjective cognitive decline. Amyloid positivity has been associated with higher levels of cognitive complaints even after accounting for depression and anxiety [15,16]. In addition, the severity of subjective cognitive complaints has been associated with higher amyloid burden [17], especially in the parietal and frontal cortices [18] and among *APOE* ε4 carriers [19]. Compared to amyloid, tau pathology is a stronger predictor of cognitive decline and progression to dementia [20–22]. However, there is limited research on the relationship between subjective cognitive complaints and tau pathology. One previous study found that subjective cognitive complaints were associated with tau aggregation, particularly in the frontal and parietal regions [23], while other studies did not find associations with tau pathology [24,25].

Our study focuses on the following three research questions: first, what is the cross-sectional association between AD neuropathology and subjective cognitive complaints in cognitively normal individuals? Second, is AD neuropathology a predictor of subjective cognitive decline over time? Third, is AD neuropathology a stronger predictor of subjective cognitive complaints compared to neuropsychological assessment-based measures of memory among cognitively normal individuals? To address these questions, we assessed the relationship of subjective cognitive complaints as measured by the Cognitive Failures Questionnaire (CFQ) with global amyloid positivity and tau PET levels in regions that are known to exhibit early tau pathology. We then compared these associations of pathology with verbal episodic memory as assessed by the California Verbal Learning Test (CVLT).

## 2 Methods

### 2.1 Participants

We used data from cognitively normal participant-visits in the neuroimaging substudy of the Baltimore Longitudinal Study of Aging (BLSA) for participants who had a ^18^F-flortaucipir (FTP) PET scan to measure phosphorylated tau burden, a ^11^C-Pittsburgh compound B (PiB) PET scan to measure fibrillar Aβ burden, completed the CFQ [26], and received the CVLT [27]. Participant selection for this analysis is detailed in the Supplement. Cognitively normal status was based on either (1) a Clinical Dementia Rating [28] of zero and ≤ 3 errors on the Blessed Information-Memory-Concentration Test [29], and therefore, the participant did not meet criteria for a consensus conference; or (2) the participant was determined to be cognitively normal based on thorough review of clinical and neuropsychological data at consensus conference. In longitudinal analyses, we included only CFQ and CVLT assessments corresponding to visits where participants were cognitively normal. Participants below the age of 60 were evaluated every 4 years, 60–79 year old participants every 2 years, and participants 80 and older annually.

Research protocols were conducted in accordance with the United States federal policy for the protection of human research subjects (45 CFR 46), approved by local Institutional Review Boards (IRB) and the National Institutes of Health, and all participants gave written informed consent at each visit. BLSA PET study was governed by the IRB of the Johns Hopkins Medical Institutions (protocol numbers NA_00051793, IRB00047185) and the BLSA study was overseen by the IRBs of the National Institute of Environmental Health Sciences and the National Institutes of Health Intramural Research Program.

### 2.2 Cognitive measures

The CFQ evaluates subjective cognitive complaints using 25 questions about the frequency of common cognitive issues within the past two weeks. We calculated total score as the sum of scores divided by four times the total number of questions answered to account for missing responses. Higher scores represent greater subjective cognitive complaints. Using the loadings by Rast et al. [30], we also calculated the three CFQ factors reflecting forgetfulness, distractibility, and false triggering as a weighted mean of the individual CFQ items (Supplement).

Verbal episodic memory was assessed by the CVLT using a single form to ensure consistency across participant-visits. We quantified immediate recall as the number of correctly remembered items across the five learning trials. A memory composite score was calculated by averaging the *z*-scores of the immediate and long delay free recall.

The Center for Epidemiological Studies–Depression (CES-D) [31] scale was used to quantify subjective depressive symptoms.

### 2.3 PET imaging

To assess phosphorylated tau, PET scans were acquired over 30 min on a Siemens High Resolution Research Tomograph (HRRT) scanner starting 75 min after an intravenous bolus injection of approximately 370 MBq of FTP. FTP PET analysis workflow is detailed in Ziontz et al. (2019) [32]. The partial volume corrected [33] 80–100 min image was used to compute standardized uptake value ratio (SUVR) images with inferior cerebellar gray matter as the reference region. We computed the average bilateral SUVR in the entorhinal cortex (EC), hippocampus, parahippocampal gyrus, and the inferior temporal gyrus (ITG). We investigated tau burden as a continuous rather than a dichotomous variable to be able to examine its subtle effects. One participant with outlying regional tau SUVRs was excluded from analyses (entorhinal SUVR of 2.0 and ITG SUVR of 2.7).

To assess fibrillar Aβ, PET scans were obtained over 70 min on either a General Electric (GE) Advance or a Siemens HRRT scanner immediately following an intravenous bolus injection of approximately 555 MBq of PiB. PiB PET analysis is detailed in Bilgel et al. (2020) [34]. HRRT scans were smoothed with a 3 mm FWHM Gaussian to match their spatial resolution to that of the GE Advance scans. Distribution volume ratio (DVR) images were computed using a spatially constrained simplified reference tissue model with cerebellar gray matter as the reference region [35]. Mean cortical Aβ burden was calculated as the average of the DVR in cingulate, frontal, parietal (including precuneus), lateral temporal, and lateral occipital cortical regions, excluding the sensorimotor strip. Leveraging longitudinal PiB PET data available on both scanners for 79 BLSA participants, we estimated the parameters of a linear model mapping mean cortical DVR values between the GE Advance and HRRT scanners and applied this mapping to all HRRT values to harmonize them with the GE Advance values. Individuals were categorized as PiB −/+ based on a mean cortical DVR threshold of 1.06, which was derived from a Gaussian mixture model fitted to harmonized values at baseline.

### 2.4 Statistical analysis

We fitted linear mixed effects models to assess the associations between longitudinal cognitive outcomes and PET biomarkers at index visit, using a separate model for CFQ and memory and per each of the four tau PET ROIs, yielding a total of 8 models. Inclusion of both amyloid status (PiB −/+) and regional tau SUVR terms allowed for the examination of their independent cross-sectional associations with the outcome (CFQ or memory), and the interactions of each of these terms with time from PET allowed for the examination of their associations with the rate of change in the CFQ and memory outcomes. We additionally included an amyloid and tau interaction to capture the synergistic cross-sectional effect of these pathologies. Age at the time of PET, age², CES-D, sex, and years of education were included as covariates. For memory models, we additionally adjusted for CVLT practice effects by including a binary term reflecting whether it was the participant’s first CVLT administration. We included a random intercept and slope for time per person. The construction of our linear mixed effects models is detailed in the Supplement. We confirmed our cross-sectional findings using linear regression models fitted using index visits only, fitted linear mixed effects models investigating each of the three CFQ factors and the CVLT measures that make up the memory composite score as outcomes, and assessed whether using a continuous index of global amyloid burden rather than binary amyloid status as a covariate affects the main results (Supplement).

Prior to fitting the models, we centered age at the time of the PET by subtracting the median age of the sample. Amyloid group was coded as −0.5 for PiB– and +0.5 for PiB+. Each cognitive outcome and continuous PET variable was standardized by subtracting its mean and dividing by its standard deviation. The means and standard deviations for this standardization were computed using a larger cross-sectional BLSA data set of cognitively normal individuals between the ages of 70 and 80, using the visit closest to age 75 per participant. As a result, the regression coefficients reported for the amyloid and tau variables are standardized coefficients.

To assess if the associations of amyloid and tau neuropathology with subjective cognitive complaints differ from their associations with verbal episodic memory, we tested the equality of the standardized regression coefficients between the CFQ and memory models using Seemingly Unrelated Regression Equations (SURE) [36]. This analysis was limited to examining cross-sectional effects and we negated CFQ to make its direction consistent with that of memory.

Linear mixed effects analyses were conducted in R version 4.3.3 [37] using lmerTest [38]. We checked the fit of all statistical models using performance [39] to ensure they were not susceptible to outliers and that there were no convergence issues. The SURE analysis was conducted using systemfit and car [40,41].

### 2.5 Data availability

Code is provided in an open repository (https://gitlab.com/bilgelm/PET-and-CFQ). BLSA data are available upon request from https://www.blsa.nih.gov after approval by the BLSA Data Sharing Proposal Review Committee.

## 3 Results

The sample consisted of 91 cognitively normal participants. Table 1 presents participant characteristics at index visit, which is defined by the requirement that they have a PiB PET within 2 years of their baseline FTP PET and a CFQ and a CVLT assessment within 6 months of their baseline FTP PET. A total of cognitively normal 241 visits were included for CFQ and 350 for CVLT (Figure 1). Pair plots of continuous variables are presented in Figures S1–S2.

**Figure 1:**
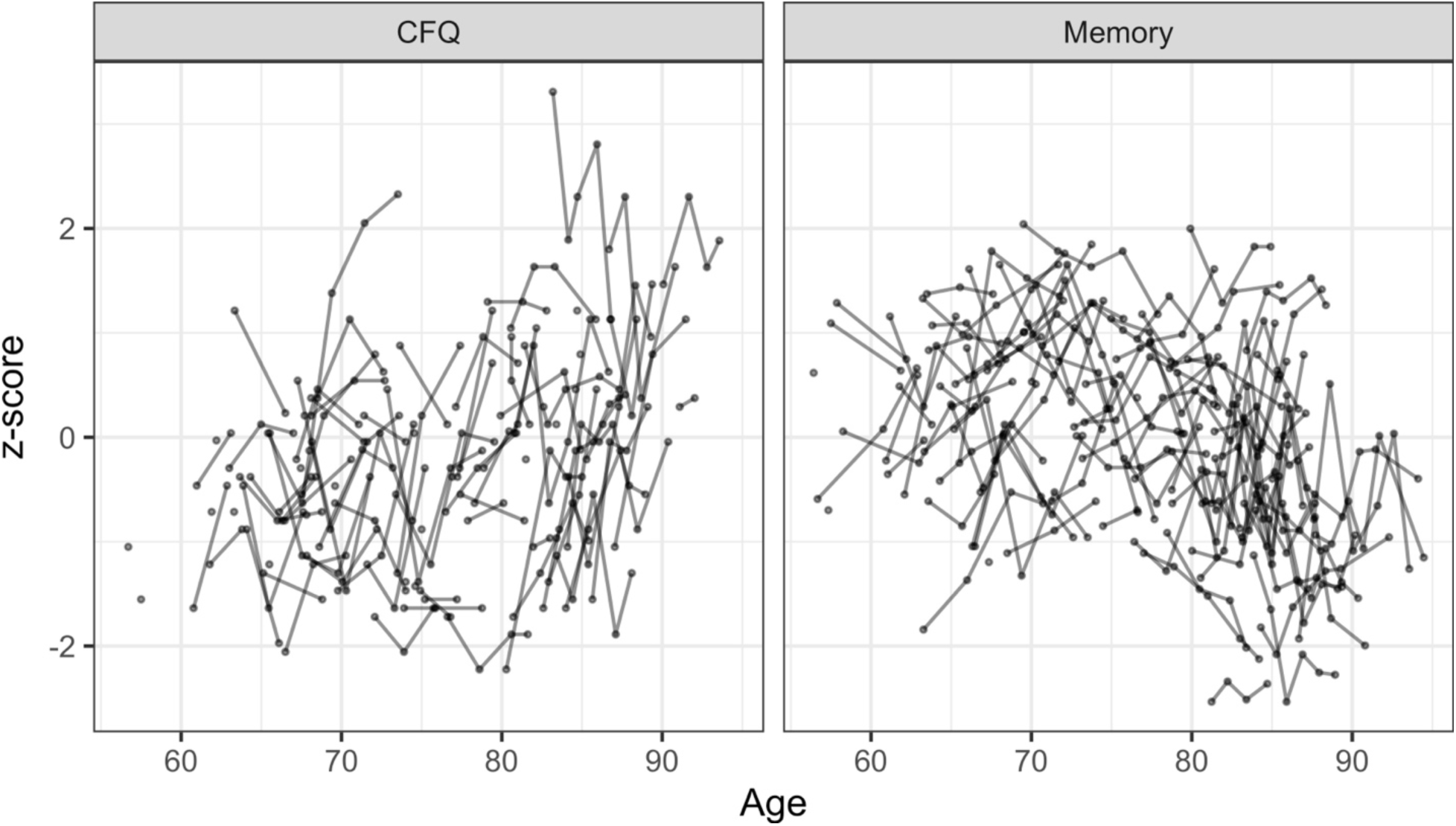
Longitudinal CFQ (left) and memory (right) scores versus age. Lower CFQ z-scores indicate lower subjective cognitive complaints and higher memory z-scores indicate better performance.

**Table 1:** Participant characteristics at index visit. For continuous and categorical variables, we report the median and interquartile range and the N and percentage, respectively.

| Characteristic | N = 91 |
| --- | --- |
| Age (yr) | 77 (69, 83) |
| Female | 52 (57%) |
| White | 70 (77%) |
| Education | 18 (16, 18) |
| *APOE* ε4+ | 30 (34%) |
| PiB+ | 24 (26%) |
| Entorhinal tau SUVR | 1.06 (0.96, 1.12) |
| Hippocampal tau SUVR | 1.29 (1.18, 1.44) |
| Parahippocampal tau SUVR | 1.01 (0.94, 1.08) |
| ITG tau SUVR | 1.22 (1.16, 1.28) |
| CES-D | 4 (1, 8) |
| CFQ score (sum of scores / 4 × # answered) | 0.28 (0.19, 0.34) |
| CFQ z-score | -0.13 (-0.88, 0.38) |
| CVLT long delay free recall | 11 (9, 13) |
| CVLT immediate recall | 52 (41, 61) |
| Memory z-score | 0.19 (-0.74, 0.77) |
| Number of CFQ visits | 3 (1, 3) |
| Number of CVLT visits | 4 (3, 5) |
| CFQ follow-up duration (yr) | 3.45 (0.00, 4.58) |
| CVLT follow-up duration (yr) | 5.06 (4.10, 6.71) |

Abbreviations: *APOE*, apolipoprotein E; CES-D, Center for Epidemiological Studies - Depression; CFQ, Cognitive Failures Questionnaire; CVLT, California Verbal Learning Test; ITG, inferior temporal gyrus; PiB, Pittsburgh compound B; SUVR, standardized uptake value ratio.

### 3.1 Subjective cognitive complaints

In adjusted linear mixed effects models, we did not find any cross-sectional associations between amyloid positivity (independent of tau) and CFQ. Amyloid positivity was related to steeper longitudinal increases in CFQ, but this effect was statistically significant only in the entorhinal tau model (estimate = 0.11, SE = 0.05, *p* = 0.041). In contrast, higher parahippocampal tau SUVR, independent of amyloid status, was associated with higher CFQ at index visit (estimate = 0.25, SE = 0.1, *p* = 0.015) (Table 2). Higher entorhinal tau SUVR at index visit was associated with with an attenuated increase in CFQ over time (estimate = −0.06, SE = 0.03, *p* = 0.047) (Figure 2). Similarly, we found a statistically significant association between higher entorhinal tau SUVR, independent of amyloid, and attenuated increase over time in the CFQ false triggering factor (Table S10), whereas the cross-sectional association with parahippocampal tau SUVR was observed only for the CFQ distractibility score (Table S9).

**Figure 2:**
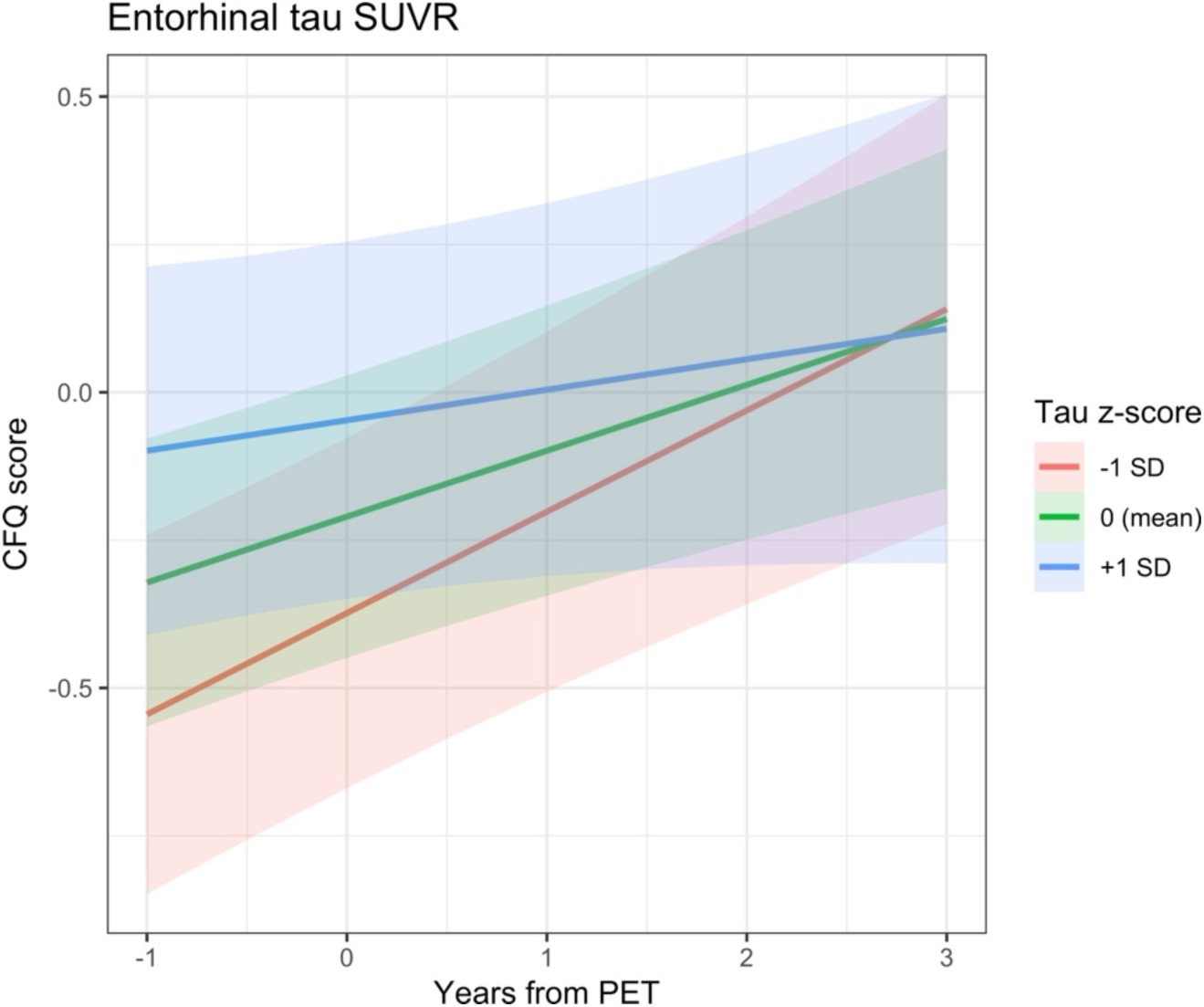
Longitudinal CFQ trajectories by entorhinal tau SUVR z-score based on linear mixed effects model estimates.

**Table 2:**
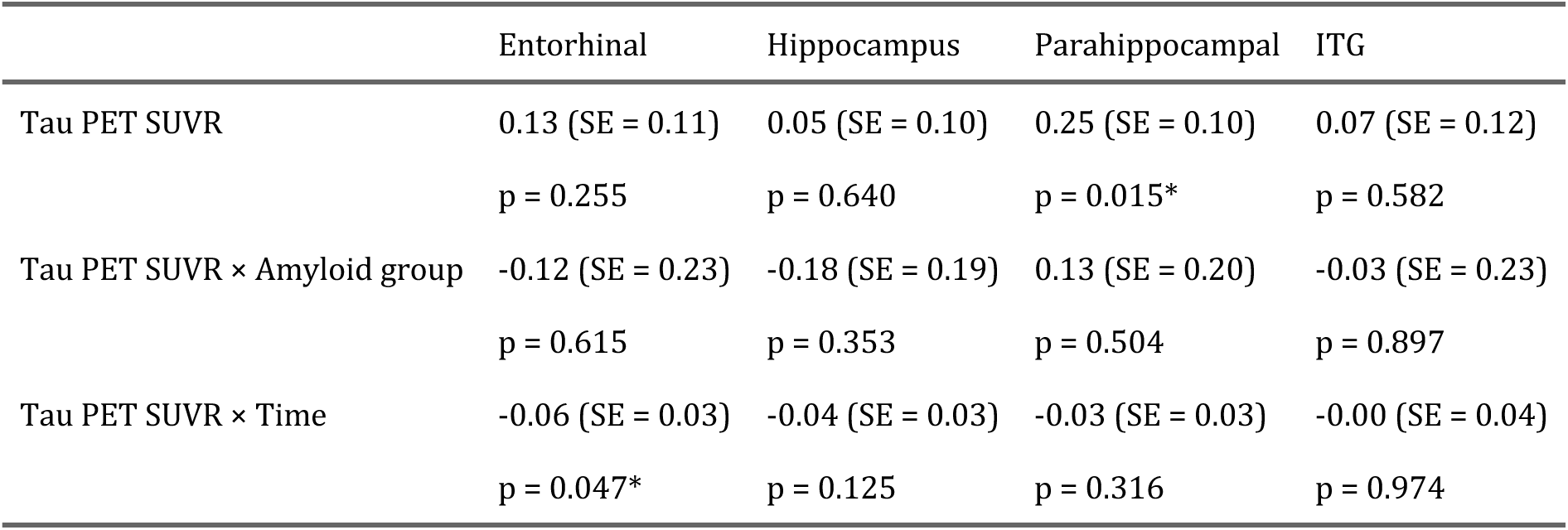
Longitudinal CFQ linear mixed effects model results. Estimated coefficients along with their standard errors (SE) and p-values for the tau PET SUVR (i.e., cross-sectional effect of tau), tau PET SUVR by amyloid group interaction (i.e., the synergistic cross-sectional effect of tau and amyloid), and the tau PET SUVR by time interaction (i.e., longitudinal effect of tau) terms in linear mixed effects models where longitudinal CFQ z-score is the outcome. Each column corresponds to a separate model. * indicates p < 0.05.

### 3.2 Verbal episodic memory

In adjusted linear mixed effects, we did not find any cross-sectional or longitudinal associations between the amyloid positivity main effect and memory. The cross-sectional associations of entorhinal and parahippocampal tau SUVR with memory were exacerbated in the presence of amyloid positivity (entorhinal tau × amyloid interaction: estimate = −0.53, SE = 0.25, *p* = 0.041; parahippocampal tau × amyloid interaction: estimate = −0.61, SE = 0.22, *p* = 0.007). Greater hippocampal tau SUVR was associated with lower concurrent memory scores (estimate = −0.27, SE = 0.11, *p* = 0.019) and a steeper decline in memory over time (estimate = −0.03, SE = 0.01, *p* = 0.040) (Table 3). We found similar cross-sectional and longitudinal associations between hippocampal tau SUVR and CVLT free recall long delay (Table S12). Greater parahippocampal tau SUVR was also associated with lower concurrent memory scores (estimate = −0.36, SE = 0.11, *p* = 0.002) and individual CVLT components (Tables S11–S12).

**Table 3:** Longitudinal memory linear mixed effects model results. Estimated coefficients along with their standard errors (SE) and p-values for the tau PET SUVR (i.e., cross-sectional effect of tau), tau PET SUVR by amyloid group interaction (i.e., the synergistic cross-sectional effect of tau and amyloid), and the tau PET SUVR by time interaction (i.e., longitudinal effect of tau) terms in linear mixed effects models where longitudinal memory z-score is the outcome. Each column corresponds to a separate model. * p < 0.05, ** p < 0.01.

|  | Entorhinal | Hippocampus | Parahippocampal | ITG |
| --- | --- | --- | --- | --- |
| Tau PET SUVR | -0.24 (SE = 0.13)<br>$p = 0.061$ | -0.27 (SE = 0.11)<br>$p = 0.019^*$ | -0.36 (SE = 0.11)<br>$p = 0.002^{**}$ | -0.17 (SE = 0.13)<br>$p = 0.198$ |
| Tau PET SUVR × Amyloid group | -0.53 (SE = 0.25)<br>$p = 0.041^*$ | -0.40 (SE = 0.21)<br>$p = 0.062$ | -0.61 (SE = 0.22)<br>$p = 0.007^{**}$ | -0.10 (SE = 0.25)<br>$p = 0.697$ |
| Tau PET SUVR × Time | -0.00 (SE = 0.02)<br>$p = 0.830$ | -0.03 (SE = 0.01)<br>$p = 0.040^*$ | -0.00 (SE = 0.01)<br>$p = 0.776$ | -0.00 (SE = 0.02)<br>$p = 0.922$ |

### 3.3 Comparison of CFQ and memory models

To examine if AD neuropathology is a stronger predictor of subjective cognitive complaints or neuropsychological assessment-based measures of memory among cognitively normal individuals, we assessed if the associations of neuropathology with subjective cognitive complaints differed from the associations of neuropathology with verbal episodic memory using Seemingly Unrelated Regression Equations (SURE). Differences between the standardized regression coefficients estimated in the CFQ and memory models were not statistically significant for amyloid group or regional tau SUVR (Table S5).

## 4 Discussion

We investigated the independent associations between PET measurements of two hallmark neuropathologies of AD and subjective cognitive complaints among cognitively normal older individuals, adjusting for age, sex, education, and depressive symptoms. Cross-sectionally, greater tau burden in the parahippocampal gyrus was associated with greater subjective cognitive complaints as assessed using the CFQ. Greater entorhinal tau SUVR was associated with an attenuated increase over a median follow-up duration of 3.4 years. In contrast, greater hippocampal SUVR was associated with a steeper decline in memory performance over a median follow-up duration of 5.1 years. When we compared models for CFQ and verbal memory directly, we did not find statistically significant differences between the associations with AD neuropathology of subjective cognitive complaints and memory scores.

The parahippocampal gyrus is part of the medial temporal lobe, a crucial region for memory processing. In agreement with our findings, several studies have also reported associations between tau in the medial temporal lobe and worse cognitive performance and cognitive decline among cognitively unimpaired individuals [42,43]. An ADNI study similarly found that self-reported complaints measured by the ECog (Measurement of Everyday Cognition) questionnaire were associated with parahippocampal tau burden as well as parietal, frontal, and global tau, although they found the strongest associations in the frontal lobe [23], whereas our study primarily analyzed regions in the temporal lobe. However, this ADNI study included those with mild cognitive impairment whereas we restricted our analysis to include only cognitively unimpaired individuals.

The association between greater entorhinal tau and attenuated increases in CFQ over time may be explained by the possibility that subjective cognitive complaints exhibit steep increases early, prior to the elevation in entorhinal tau burden, with the rate of increase in subjective cognitive complaints tapering off later as entorhinal tau burden increases (Figure 3). This possible explanation accounts for both the cross-sectional associations we observed, with higher tau burden being associated with greater subjective cognitive complaints, and the association of higher tau burden with attenuated increases in subjective cognitive complaints. Another possible explanation for this association is that as an individual experiences advancing pathology and presumably poorer objective memory, they may experience a lack of insight into their cognitive deficits, resulting in a greater denial of symptoms. Larger studies examining both subjective cognitive complaints and tau burden longitudinally will be needed to test these hypotheses.

**Figure 3:**
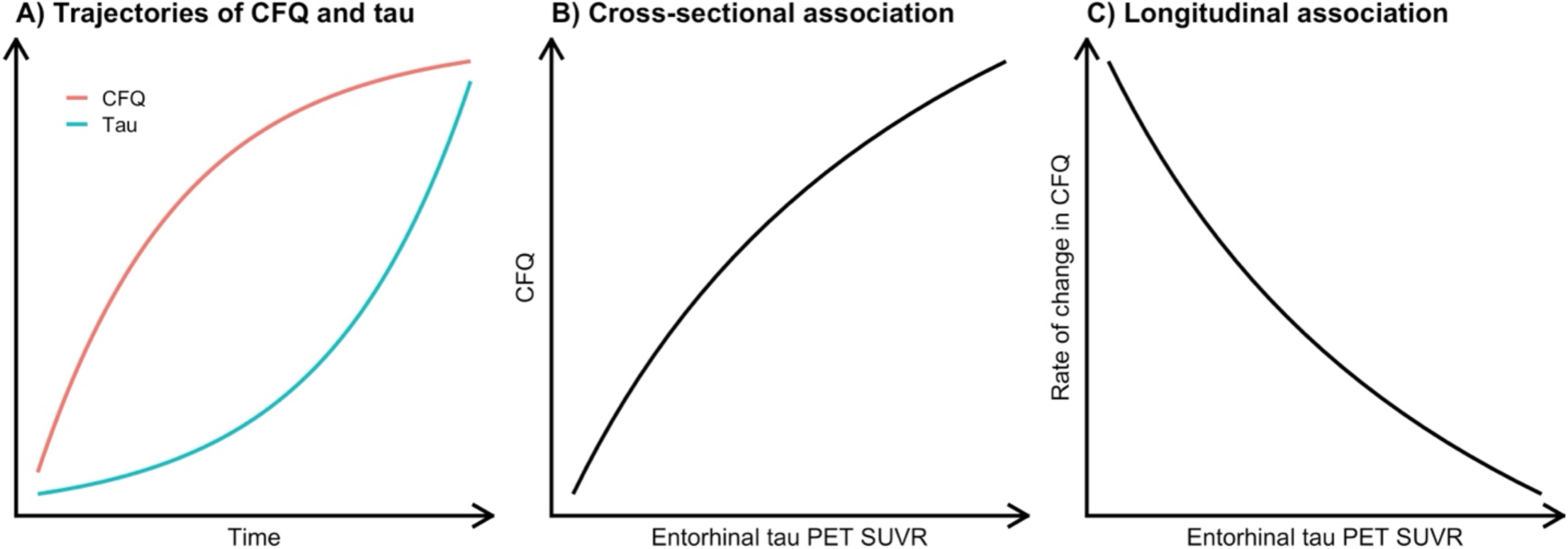
A possible explanation of the observed cross-sectional and longitudinal associations between tau PET SUVR and CFQ. The trajectories shown in panel A for tau (blue) and CFQ (red) yield a positive cross-sectional association between tau and CFQ as shown in panel B and a negative association between tau and rate of change in CFQ as shown in panel C.

Compared to the total CFQ score, the CFQ false triggering factor, but not forgetfulness or distractibility, had a statistically significant longitudinal association of similar size with entorhinal tau, indicating that our longitudinal finding may be driven by this factor [30].

Our findings differ from previous reports in several ways. First, a Harvard Aging Brain Study found that entorhinal tau PET was associated cross-sectionally with subjective cognitive decline as measured by the Memory Functioning Questionnaire, Everyday Cognition battery, and a 7-item questionnaire [44]. While our cross-sectional results are consistent with this finding, our results did not reach statistical significance. In addition, we did not find a relationship between amyloid positivity and longitudinal change in memory, in contrast to longitudinal studies that have consistently indicated this association [45,46], including from the BLSA [47]. This is likely attributable to our inclusion of participants who remained cognitively normal throughout the longitudinal follow-up period included in these analyses and our limited sample size. Because we restricted the sample to cognitively normal participants, the effect of amyloid and tau on verbal memory decline was likely subtle, therefore requiring a larger sample size to detect a statistically significant association. We note, however, that the effect we estimated was in the expected direction. Additionally, the present study adjusted for regional tau pathology in the amyloid models, whereas the previous analysis in the BLSA did not. Second, unlike a previous BLSA analysis that found that elevated baseline entorhinal tau was linked to steeper memory decline prior to tau PET [32], we did not find an association between entorhinal tau and rate of memory decline. This difference may be attributed to the extent of CVLT follow-up in the present study compared with the Ziontz et al. study, as we limited the inclusion of CVLT scores to within five years of their index tau PET visit. In our models, amyloid and tau pathologies were assumed to have additive effects on the outcomes we investigated, but research suggests that their effects might be synergistic, with individuals having both pathologies exhibiting steeper cognitive decline. However, we were unable to investigate this synergistic effect due to our limited sample size.

We did not adjust our results for multiple comparisons across the four *a priori* selected brain regions we investigated for tau pathology. At a type I error rate of α = 0.05, our family-wise error rate (FWER) is bounded above by 1 – (1 – 0.05)^4^ ≈ 18.5%, which is the FWER under the assumption of independence across hypotheses. Given that these four brain regions exhibit moderate to high correlations in their tau SUVRs (Figure S2), the true FWER of our analyses is much lower than this upper bound. Our results should be confirmed in larger samples with sufficient statistical power that enables the implementation of a multiple comparison correction framework.

This study has limitations. Our study accounted for certain factors that are associated with subjective cognitive complaints, such as severe psychiatric or neurological disorders, which are exclusion criteria for the BLSA, and depressive symptoms, which we adjusted for using CES-D as a covariate. However, there may be other conditions that affect subjective cognitive complaints that we did not account for, such as personality, medication use, substance use, and cultural background. For instance, one study found that CFQ was more associated with personality domains, such as conscientiousness and neuroticism, rather than objective cognitive performance [10]. In addition, our PET measures, particularly in the hippocampus, may be confounded by spill-over of non-specific binding signal in the choroid plexus. Lastly, the BLSA cohort is a highly educated, healthy sample largely comprising White participants. Therefore, it is uncertain how these findings will generalize to a more socioeconomically diverse cohort.

Our study also has important strengths. This study extensively characterized participants through our available longitudinal CFQ measures. We made a direct comparison between subjective and objective measures by investigating associations of tau pathology with CFQ and CVLT. We also created a proportion variable to include those who had missing items on the questionnaire to maximize our sample size. In addition, we used subjective cognitive complaints as a continuous variable, unlike other studies that used subjective cognitive measures to identify participants as part of a subjective cognitive impairment group and then compared to healthy, MCI, and AD groups.

In conclusion, these findings add to our growing knowledge of the relationship between AD pathology and cognition. A previous BLSA study found that subjective cognitive complaints, also measured by the CFQ, were predictive of declining cognitive performance, particularly in verbal memory [6]. Other studies have found that those with subjective memory complaints combined with baseline AD neuropathology may be more likely to develop dementia and cognitive decline subsequently [48,49]. Therefore, our findings support the utility of including measures of subjective cognitive assessments as early indicators for AD. Early amyloid and tau accumulation may have cognitive effects that could be reflected in subjective cognitive questionnaires such as the CFQ. Future studies investigating the relationship between AD neuropathology and subjective cognitive complaints, particularly among larger and more diverse cohorts, can help clinicians detect those at risk for developing AD at the earliest opportunity and develop targeted interventions.

## Acknowledgments

We are grateful to the BLSA participants and staff for their dedication to these studies. We thank Dr. Andrea Shafer for her statistical suggestions in the early stages of our analyses. This research was supported by the Intramural Research Program of the National Institute on Aging, NIH.

## Consent Statement

All human subjects involved in this study provided informed consent prior to participation.

## Conflicts of Interest

The authors of this study declare that there are no conflicts of interest related to this work.

## Supplementary Material

### Participant and visit selection

Enrollment criteria for the BLSA PET neuroimaging substudy included having no evidence of central nervous system disease, severe cardiovascular or pulmonary disease, major psychiatric disorders, or metastatic cancer.

For inclusion in these analyses, participants needed to have a PiB PET within 2 years of their first FTP PET as well as a CFQ and a CVLT assessment within 6 months of their baseline FTP or PiB PET. CFQ assessments where participants did not answer more than half of the questions on the CFQ (i.e., missed the back of the questionnaire sheet) were excluded. Of 113 participants who underwent both amyloid and tau PET imaging, 103 had at least one cognitively normal PET visit, with the two PET scans being ≤ 2 years apart. 101 of these participants also had at least one visit with a CES-D assessment within 2 weeks of PET. Of these, 91 had at least one visit where they had both CFQ and CVLT assessments within 6 months of PET.

We examined all CFQ and CVLT assessments up to 5 years prior to and following the baseline FTP PET scan. All CFQ and CVLT measurements were obtained from 2011 to 2022.

### Pair plots of cross-sectional continuous variables

Cross-sectionally, CFQ was inversely correlated with memory (*r* = −0.4, *p* < 0.001) (Figure S1). CFQ was positively correlated with age (*r* = 0.45, *p* < 0.001) and CES-D (*r* = 0.51, *p* < 0.001). Memory was negatively correlated with age (*r* = −0.36, *p* < 0.001) and CES-D (*r* = −0.32, *p* = 0.002). We did not find statistically significant correlations of CFQ or memory with years of education.

**Figure S1:**
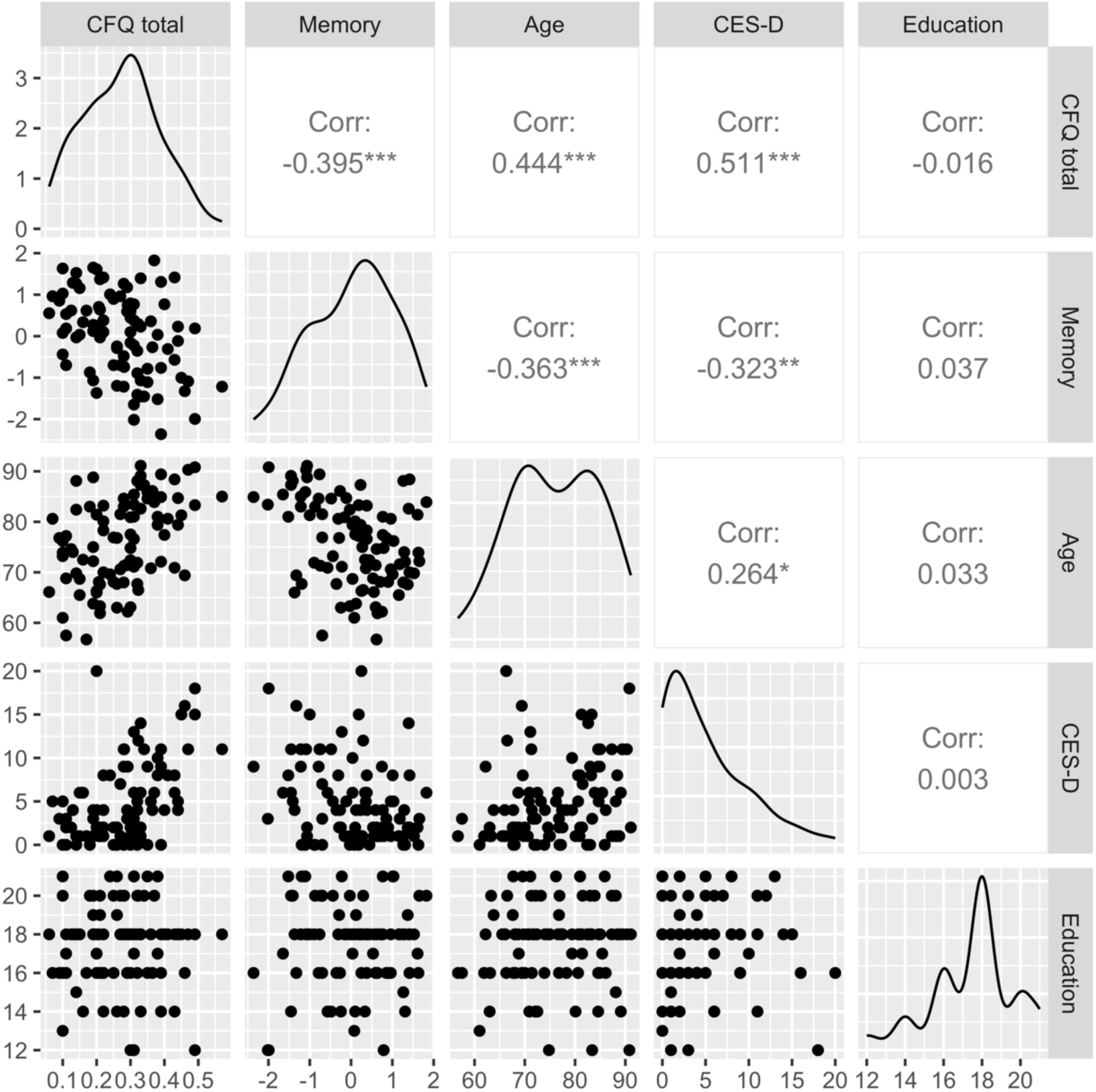
Pair plots of CFQ, memory, age, CES-D, and years of education. * < 0.05, ** < 0.01, *** < 0.001.

**Figure S2:**
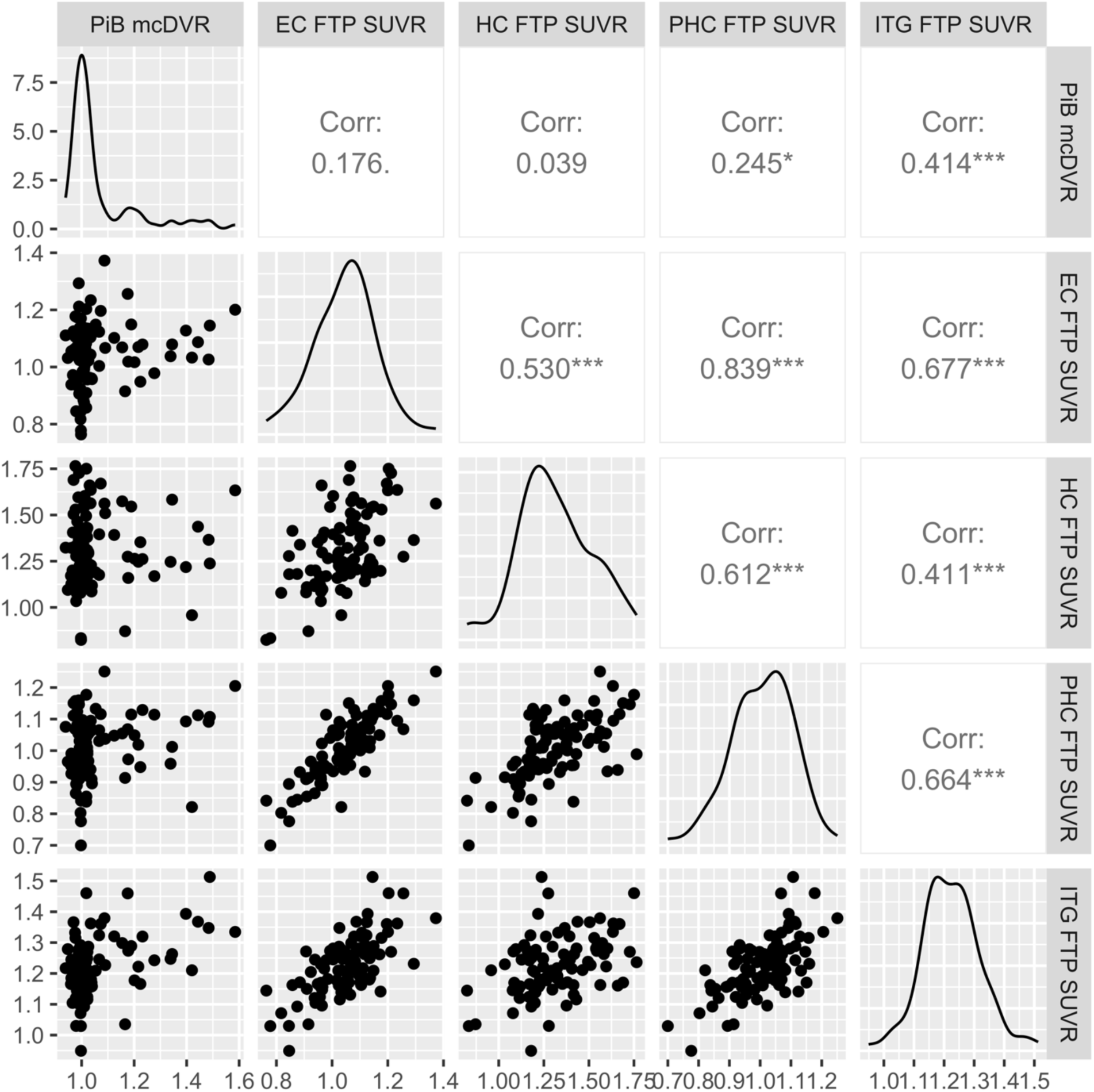
Pair plots of mean cortical PiB DVR and regional FTP SUVR. EC, entorhinal cortex; FTP, flortaucipir; HC, hippocampus; ITG, inferior temporal gyrus; mcDVR, mean cortical distribution volume ratio; PHC, parahippocampus; PiB = Pittsburgh compound B. * < 0.05, *** < 0.001.

### z-scoring

#### CFQ scores

For 223 out of the 241 CFQ assessments, participants answered all 25 CFQ questions. 17 assessments had one unanswered CFQ question and 1 assessment had two unanswered CFQ questions, with questions 15 and 20 being the most likely ones to be unanswered.

The CFQ total score variable was *z*-scored using the mean (0.3) and standard deviation (0.12) computed using a cross-sectional BLSA data set of 147 cognitively normal individuals between the ages of 70 and 80, using the visit closest to age 75 per participant.

#### Verbal episodic memory

For *z*-scoring the CVLT long delay free recall and immediate recall variables, we used mean = 10.7 (SD = 3.3) and mean = 51.3 (SD = 11.6), respectively, which were computed using a cross-sectional BLSA data set of 957 cognitively normal individuals between the ages of 70 and 80, using the visit closest to age 75 per participant.

### Longitudinal characterization of CFQ and memory scores

In addition to visualizing longitudinal trajectories of CFQ and memory scores as a function of age (Figure 1), we also visualized them as a function of time from PET (Figure S3).

**Figure S3:**
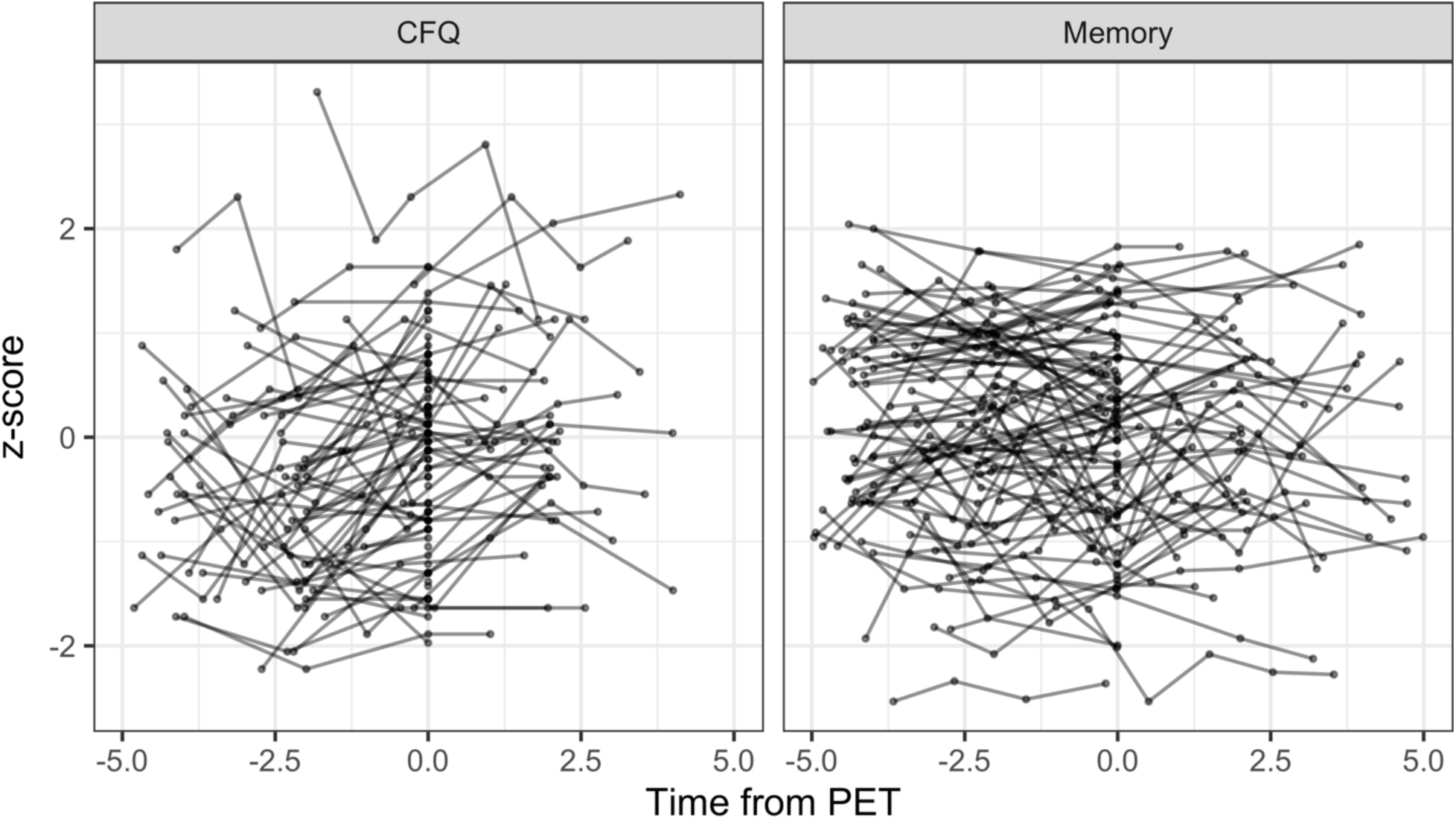
Longitudinal CFQ (left) and memory (right) scores versus time from PET. Lower CFQ z-scores indicate lower subjective cognitive complaints and higher memory z-scores indicate better performance.

To construct our linear mixed effects models, we employed a multi-stage process where we first examined models without AD pathology, then models with only amyloid or only tau pathology, and finally models with both amyloid and tau pathologies. The linear mixed effects model results for the intermediate models are presented in this subsection.

### Without AD neuropathology variables

First, we conducted analyses that did not include amyloid or tau pathology as covariates to examine the longitudinal changes in CFQ total score and memory *z*-score.

**Table S1:** Estimated coefficients along with their standard errors (SE) and p-values in linear mixed effects models where longitudinal CFQ (left) or longitudinal memory (right) z-score is the outcome. Each column corresponds to a separate model. * p < 0.05, ** p < 0.01, *** p < 0.001.

|  | CFQ | Memory |
| --- | --- | --- |
| Intercept | -0.32 (SE = 0.11)<br>p = 0.003** | 0.19 (SE = 0.17)<br>p = 0.247 |
| Age at index visit | 0.03 (SE = 0.01)<br>p = <0.001*** | -0.04 (SE = 0.01)<br>p = <0.001*** |
| Age<sup>2</sup> | 0.00 (SE = 0.00)<br>p = 0.402 | -0.00 (SE = 0.00)<br>p = 0.019* |
| Sex (ref = Female) | -0.06 (SE = 0.16)<br>p = 0.728 | 0.09 (SE = 0.18)<br>p = 0.603 |
| Education | -0.00 (SE = 0.04)<br>p = 0.996 | 0.00 (SE = 0.04)<br>p = 0.986 |
| CES-D | 0.09 (SE = 0.02)<br>p = <0.001*** | -0.03 (SE = 0.02)<br>p = 0.093 |
| Practice effect |  | 0.11 (SE = 0.12)<br>p = 0.357 |
| Time | 0.10 (SE = 0.02)<br>p = <0.001*** | 0.00 (SE = 0.01)<br>p = 0.972 |

### With amyloid pathology only

Next, we conducted an analysis adjusting for only amyloid (but not tau) pathology.

**Table S2:** Estimated coefficients along with their standard errors (SE) and p-values in linear mixed effects models where longitudinal CFQ (left) or longitudinal memory (right) z-score is the outcome. Each column corresponds to a separate model. * p < 0.05, ** p < 0.01, *** p < 0.001.

|  | CFQ | Memory |
| --- | --- | --- |
| Intercept | -0.34 (SE = 0.11)<br>p = 0.003** | 0.20 (SE = 0.17)<br>p = 0.253 |
| Age at index visit | 0.03 (SE = 0.01)<br>p = <0.001*** | -0.04 (SE = 0.01)<br>p = <0.001*** |
| Age<sup>2</sup> | 0.00 (SE = 0.00)<br>p = 0.501 | -0.00 (SE = 0.00)<br>p = 0.030* |
| Sex (ref = Female) | -0.07 (SE = 0.16)<br>p = 0.661 | 0.11 (SE = 0.18)<br>p = 0.534 |
| Education | -0.00 (SE = 0.04)<br>p = 0.899 | 0.01 (SE = 0.04)<br>p = 0.832 |
| CES-D | 0.09 (SE = 0.02)<br>p = <0.001*** | -0.03 (SE = 0.02)<br>p = 0.083 |
| Practice effect |  | 0.12 (SE = 0.12)<br>p = 0.293 |
| Amyloid group (ref = PiB-) | -0.09 (SE = 0.18)<br>p = 0.625 | 0.10 (SE = 0.21)<br>p = 0.636 |
| Time | 0.12 (SE = 0.03)<br>p = <0.001*** | -0.01 (SE = 0.02)<br>p = 0.555 |
| Amyloid group × Time | 0.10 (SE = 0.05)<br>p = 0.070 | -0.04 (SE = 0.03)<br>p = 0.177 |

### With tau pathology only

Third, we conducted an analysis adjusting for only tau (but not amyloid) pathology.

**Table S3:**
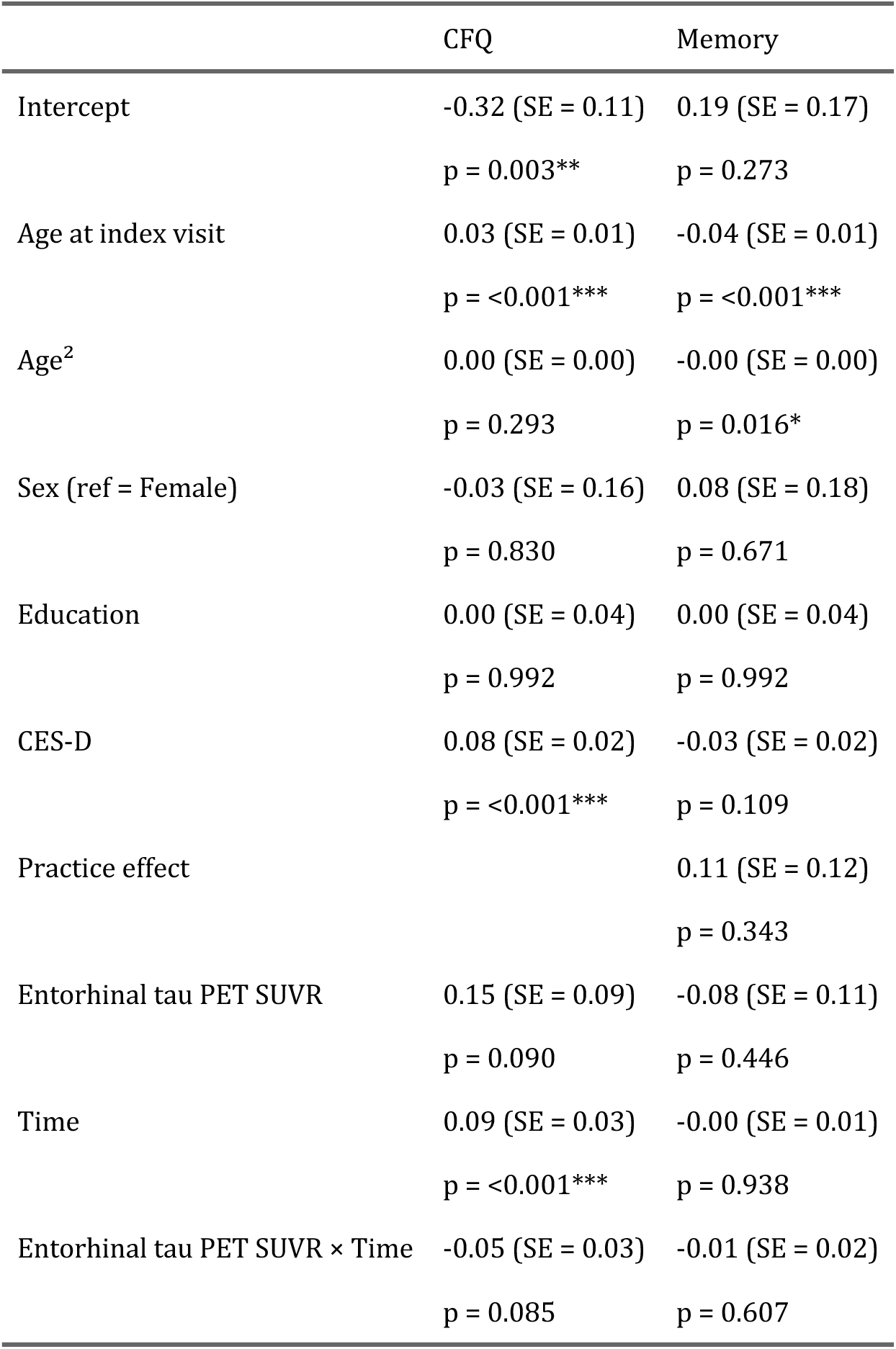
Estimated coefficients along with their standard errors (SE) and p-values in linear mixed effects models where longitudinal CFQ (left) or longitudinal memory (right) z-score is the outcome. Each column corresponds to a separate model. * p < 0.05, ** p < 0.01, *** p < 0.001.

### With both amyloid and tau pathology

Finally, we included both amyloid and tau pathology in the models and investigated whether interactions between the two pathologies should be retained. We conducted analyses including the interaction between amyloid and tau (amyloid × tau to examine cross-sectional interactions and amyloid × tau × time to examine associations with longitudinal rates of change in CFQ and memory scores). We did not observe any statistically significant amyloid × tau interactions in association with CFQ, but there were statistically significant associations with memory. We did not find any statistically significant amyloid × tau × time interactions in either the CFQ (estimate = 0, SE = 0.07, *p* = 0.985) or the memory model (estimate = 0.01, SE = 0.04, *p* = 0.740), so this three-way interaction term was not included in the final models.

For CFQ, the model that included both amyloid and tau (i.e., amyloid, amyloid × time, tau, tau × time, and amyloid × tau terms; log likelihood = −260, AIC = 546) yielded a better fit than the model with amyloid only (log likelihood = −256, AIC = 544; likelihood ratio test *p* = 0.04). For memory, the model with both amyloid and tau (log likelihood = −327, AIC = 682) had a better fit than the model with tau only (log likelihood = −323, AIC = 681; likelihood ratio test *p* = 0.06).

Based on these findings, the final models included both amyloid and tau pathology, their interactions with time, and a cross-sectional amyloid × tau term. The estimated coefficients for the final CFQ and memory models are presented for the entorhinal area in Table S4. Figure S4 illustrates the observed and fitted longitudinal CFQ trajectories.

**Table S4:**
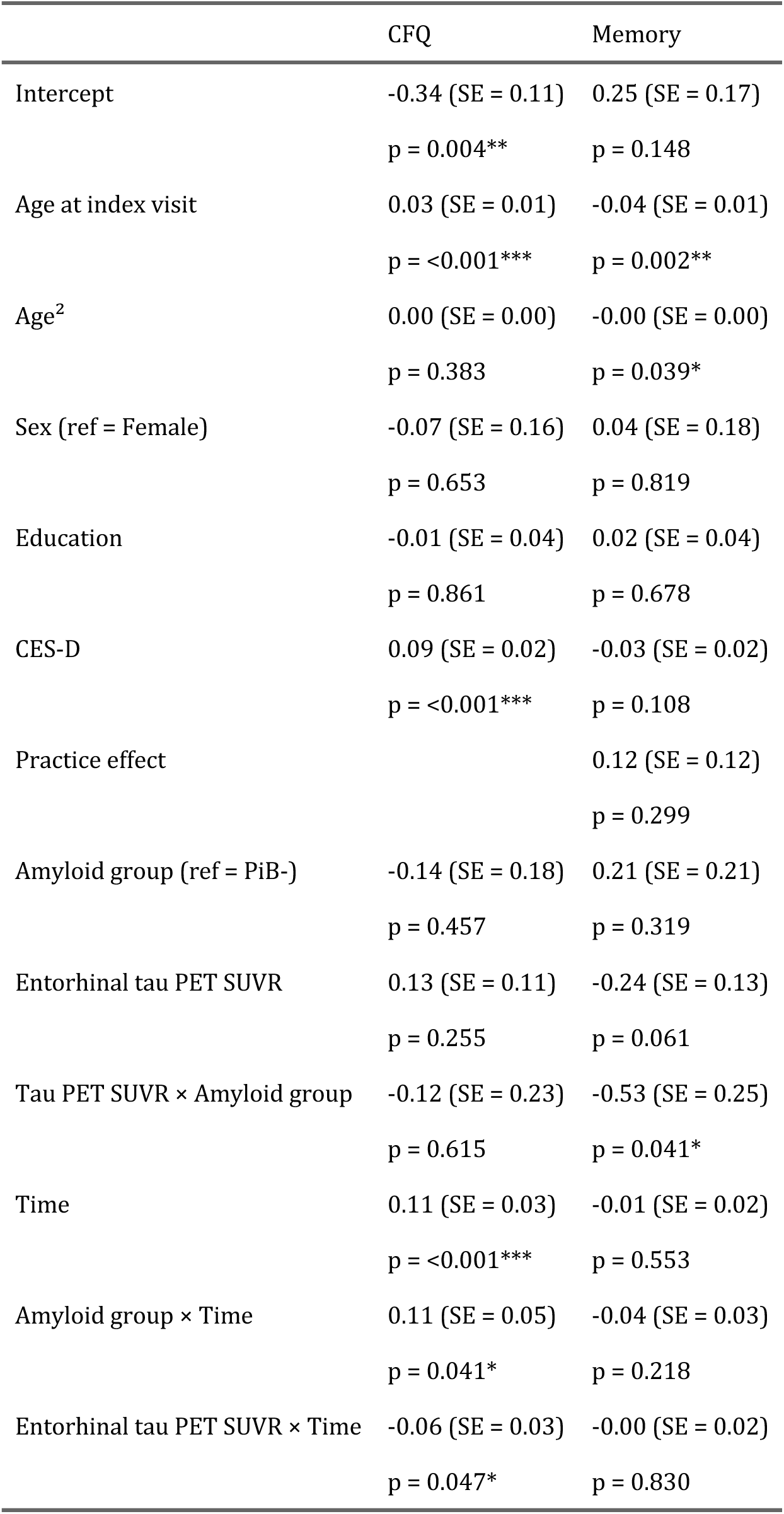
Estimated coefficients along with their standard errors (SE) and p-values in linear mixed effects models where longitudinal CFQ (left) or longitudinal memory (right) z-score is the outcome. Each column is a separate model. * p < 0.05, ** p < 0.01, *** p < 0.001.

**Figure S4:**
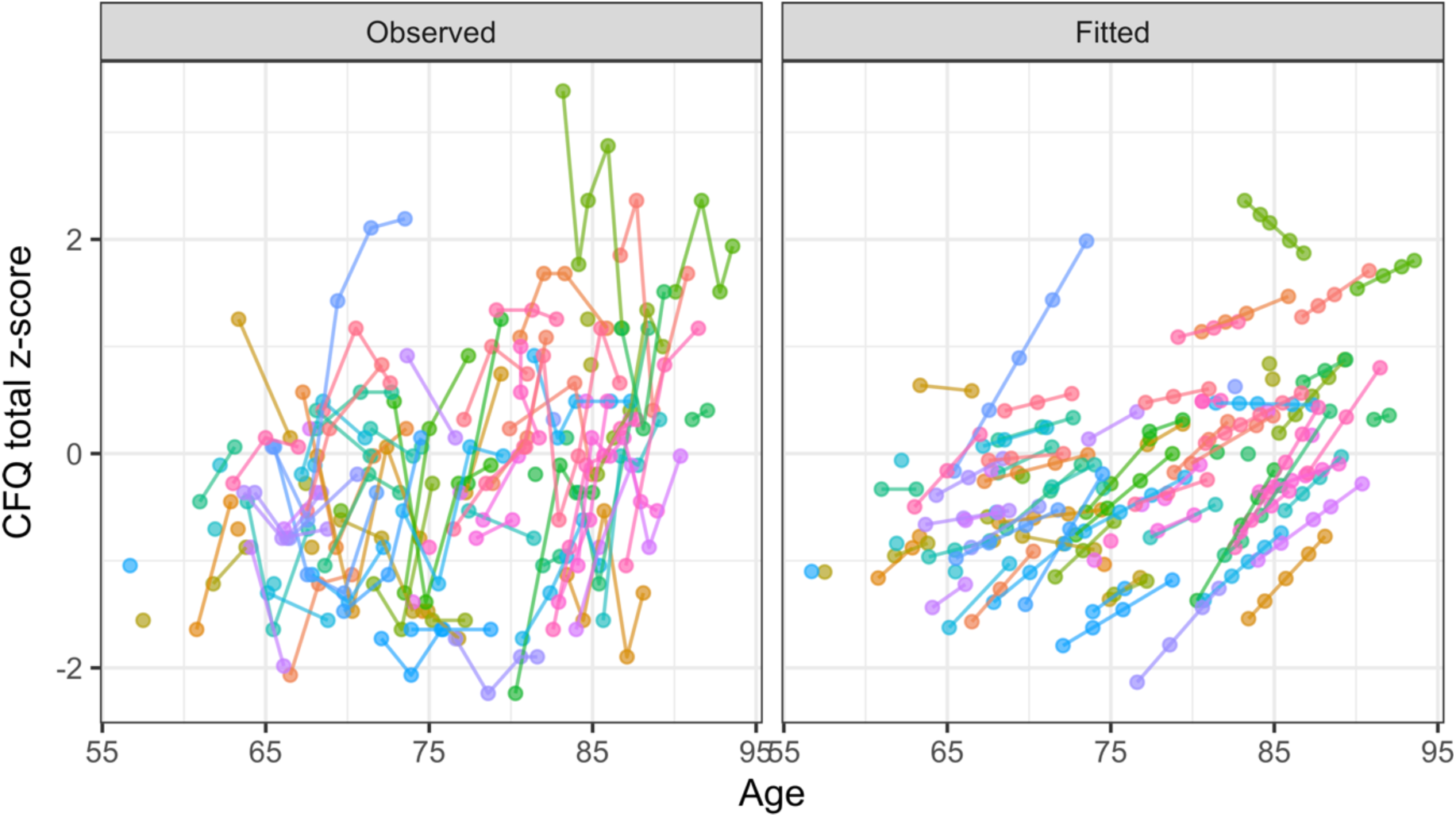
Observed (left) and fitted (right) longitudinal CFQ total z-score trajectories using final linear mixed effects models.

### Results of the SURE analysis

**Table S5:** Seemingly Unrelated Regression Equations (SURE) analysis results. The term column reflects the linear regression coefficient whose equality between the CFQ and memory models is being tested.

| Term | Entorhinal | Hippocampus | Parahippocampal | ITG |
| --- | --- | --- | --- | --- |
| Amyloid group | F = 0; p = 0.98 | F = 0.02; p = 0.89 | F = 0; p = 0.96 | F = 0; p = 0.97 |
| Tau SUVR | F = 0.04; p = 0.84 | F = 0.13; p = 0.72 | F = 0; p = 0.99 | F = 0.05; p = 0.82 |

### Linear regression models

Linear regression, using index visits only, was used to confirm our cross-sectional associations in the linear mixed effects models. Most of the estimates were similar to those based on the linear mixed effects models. We replicated the statistical significances for the cross-sectional associations between parahippocampal tau and CFQ, parahippocampal tau and memory, and the parahippocampal tau × amyloid group interaction on memory.

#### Subjective cognitive complaints

**Figure S5:**
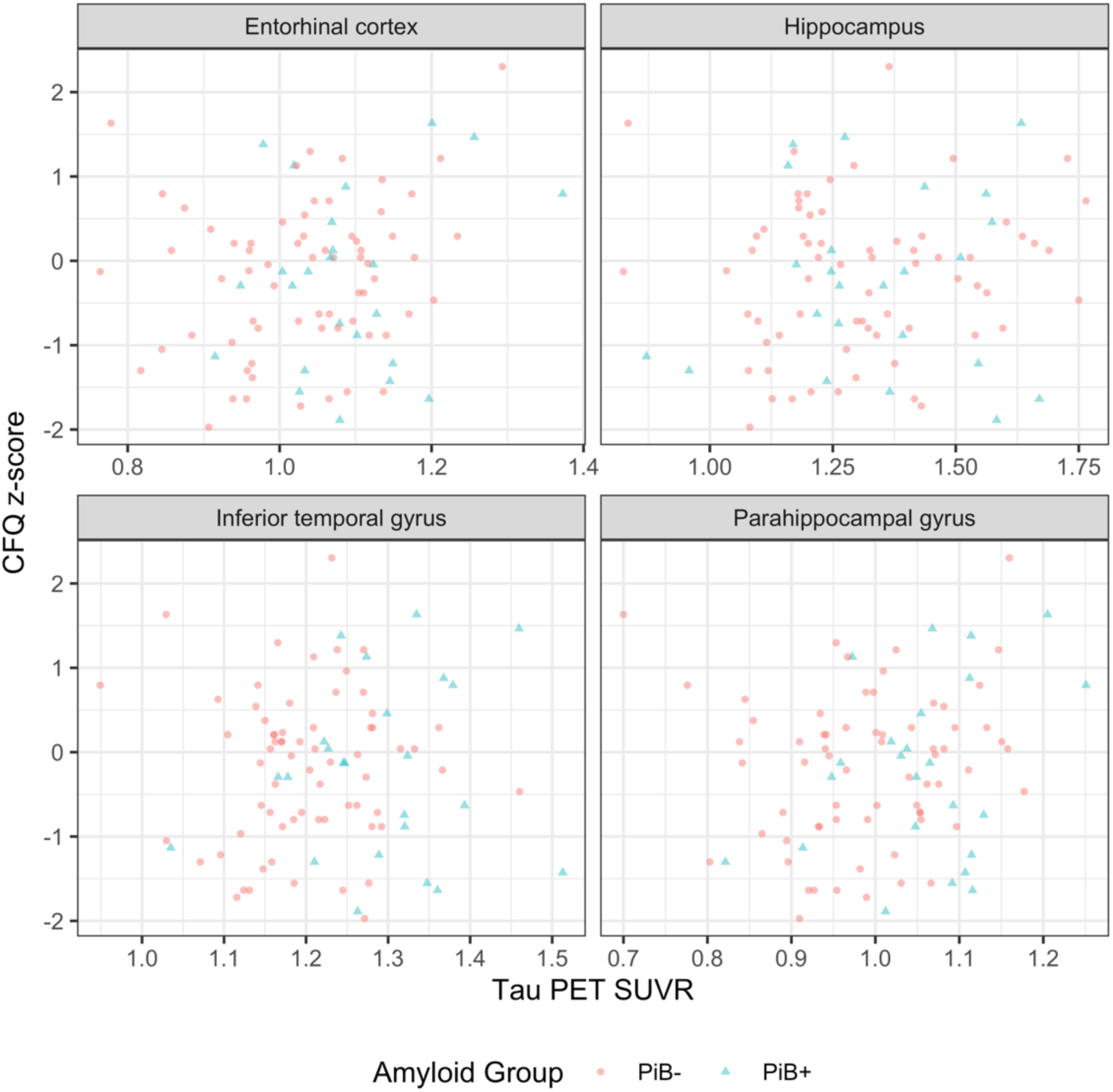
CFQ score at index visit versus regional tau SUVR.

**Table S6:** Linear regression coefficients, their standard errors, and p-values for the tau PET SUVR term for models where CFQ is the outcome. Each column corresponds to a separate model. ** p < 0.01.

|  | Entorhinal | Hippocampus | Parahippocampal | ITG |
| --- | --- | --- | --- | --- |
| Tau PET SUVR | 0.12 (SE = 0.12)<br>p = 0.344 | 0.10 (SE = 0.11)<br>p = 0.354 | 0.29 (SE = 0.11)<br>p = 0.010** | 0.06 (SE = 0.13)<br>p = 0.627 |
| Tau PET SUVR × Amyloid group | -0.20 (SE = 0.24)<br>p = 0.414 | -0.12 (SE = 0.21)<br>p = 0.552 | 0.11 (SE = 0.21)<br>p = 0.605 | -0.06 (SE = 0.24)<br>p = 0.814 |

#### Verbal episodic memory

**Figure S6:**
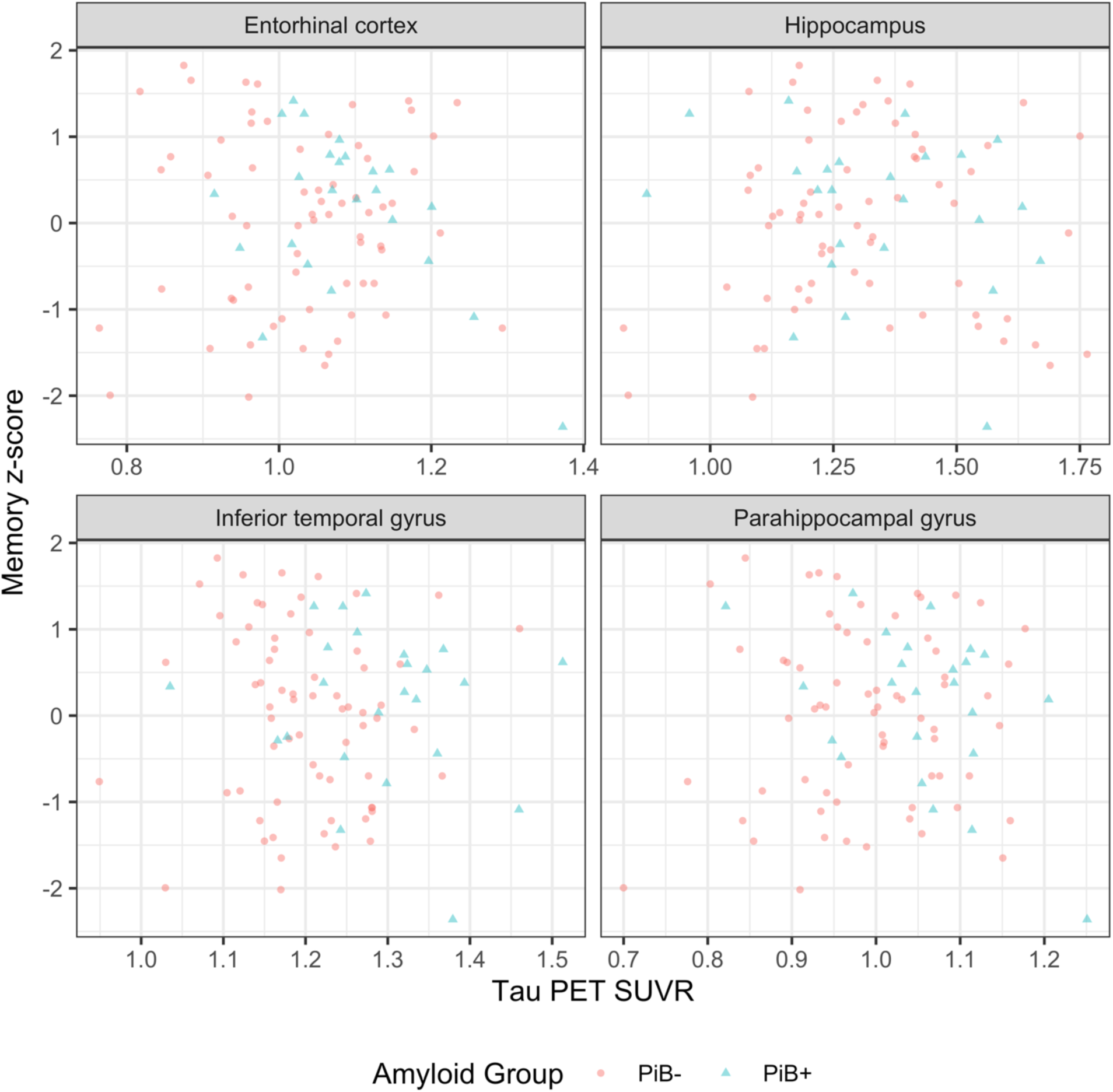
Memory z-score at index visit versus regional tau SUVR.

**Table S7:** Linear regression coefficients, their standard errors, and p-values for the tau PET SUVR term for models where the outcome is the memory z-score. Each column corresponds to a separate model. * indicates p < 0.05.

|  | Entorhinal | Hippocampus | Parahippocampal | ITG |
| --- | --- | --- | --- | --- |
| Tau PET SUVR | -0.15 (SE = 0.14)<br>p = 0.299 | -0.15 (SE = 0.12)<br>p = 0.232 | -0.28 (SE = 0.13)<br>p = 0.026* | -0.10 (SE = 0.15)<br>p = 0.501 |
| Tau PET SUVR × Amyloid group | -0.33 (SE = 0.28)<br>p = 0.254 | -0.25 (SE = 0.24)<br>p = 0.289 | -0.50 (SE = 0.25)<br>p = 0.050* | 0.07 (SE = 0.28)<br>p = 0.813 |

### Other linear mixed effects models

#### Subjective cognitive complaints

Using the loadings reported by Rast et al. [30], we calculated the three CFQ factor scores reflecting forgetfulness, distractibility, and false triggering as a weighted mean of the individual CFQ scores. For assessments with unanswered questions, the missing questions (and their corresponding weights) were excluded when computing the weighted mean. The distractibility factor is defined by Rast et al. as “being absentminded or easily disturbed in one’s focused attention in social situations,” and comprises loadings from CFQ items such as “Do you fail to hear people speaking to you when you are doing something else?” and “Do you leave important letters unanswered for days?” The false triggering factor is defined as the “interrupted processing of sequences of cognitive and motor actions,” and comprises loadings from CFQ items such as “Do you find you forget why you went from one part of the house to the other?” and “Do you find you forget what you went to the store to buy?” The forgetfulness factor is defined as “a tendency to let go from one’s mind something known or planned” and comprises loadings from CFQ items such as “Do you find you forget people’s names” and “Do you bump into people?” [30].

The forgetfulness, distractibility, and false triggering factor scores were based on questions without any missingness for 230, 233, and 226 CFQ assessments, respectively.

For z-scoring the forgetfulness, distractibility, and false triggering factor scores, we used mean = 1.44 (SD = 0.52), mean = 1.11 (SD = 0.49), and mean = 1.14 (SD = 0.49), respectively.

We repeated the linear mixed effects models using each of the CFQ factors (forgetfulness, distractibility, and false triggering) rather than the total CFQ score as the outcome.

**Table S8:** Estimated coefficients along with their standard errors (SE) and p-values for the tau PET SUVR (i.e., cross-sectional effect) and the tau PET SUVR by time interaction (i.e., longitudinal effect) terms in linear mixed effects models where longitudinal CFQ forgetfulness is the outcome. Each column corresponds to a separate model.

|  | Entorhinal | Hippocampus | Parahippocampal | ITG |
| --- | --- | --- | --- | --- |
| Tau PET SUVR | 0.07 (SE = 0.12)<br>p = 0.563 | 0.04 (SE = 0.10)<br>p = 0.659 | 0.17 (SE = 0.10)<br>p = 0.109 | 0.06 (SE = 0.12)<br>p = 0.597 |
| Tau PET SUVR × Amyloid group | -0.24 (SE = 0.24)<br>p = 0.311 | -0.22 (SE = 0.19)<br>p = 0.254 | -0.01 (SE = 0.21)<br>p = 0.954 | -0.10 (SE = 0.23)<br>p = 0.656 |
| Tau PET SUVR × Time | -0.06 (SE = 0.03)<br>p = 0.067 | -0.05 (SE = 0.03)<br>p = 0.059 | -0.04 (SE = 0.03)<br>p = 0.199 | -0.01 (SE = 0.04)<br>p = 0.733 |

**Table S9:** Estimated coefficients along with their standard errors (SE) and p-values for the tau PET SUVR (i.e., cross-sectional effect) and the tau PET SUVR by time interaction (i.e., longitudinal effect) terms in linear mixed effects models where longitudinal CFQ distractibility is the outcome. Each column corresponds to a separate model. ** < 0.01.

|  | Entorhinal | Hippocampus | Parahippocampal | ITG |
| --- | --- | --- | --- | --- |
| Tau PET SUVR | 0.17 (SE = 0.12)<br>p = 0.157 | 0.05 (SE = 0.10)<br>p = 0.605 | 0.27 (SE = 0.10)<br>p = 0.011* | 0.11 (SE = 0.12)<br>p = 0.379 |
| Tau PET SUVR × Amyloid group | -0.05 (SE = 0.24)<br>p = 0.832 | -0.20 (SE = 0.20)<br>p = 0.307 | 0.14 (SE = 0.21)<br>p = 0.501 | 0.01 (SE = 0.24)<br>p = 0.954 |
| Tau PET SUVR × Time | -0.04 (SE = 0.03)<br>p = 0.137 | -0.02 (SE = 0.02)<br>p = 0.289 | -0.01 (SE = 0.02)<br>p = 0.587 | 0.01 (SE = 0.04)<br>p = 0.742 |

**Table S10:** Estimated coefficients along with their standard errors (SE) and p-values for the tau PET SUVR (i.e., cross-sectional effect) and the tau PET SUVR by time interaction (i.e., longitudinal effect) terms in linear mixed effects models where longitudinal CFQ false triggering is the outcome. Each column corresponds to a separate model. * p < 0.05.

|  | Entorhinal | Hippocampus | Parahippocampal | ITG |
| --- | --- | --- | --- | --- |
| Tau PET SUVR | 0.09 (SE = 0.11)<br>p = 0.422 | -0.00 (SE = 0.10)<br>p = 0.999 | 0.19 (SE = 0.10)<br>p = 0.054 | 0.02 (SE = 0.11)<br>p = 0.830 |
| Tau PET SUVR × Amyloid group | -0.14 (SE = 0.22)<br>p = 0.526 | -0.17 (SE = 0.18)<br>p = 0.352 | 0.11 (SE = 0.20)<br>p = 0.587 | -0.01 (SE = 0.22)<br>p = 0.960 |
| Tau PET SUVR × Time | -0.07 (SE = 0.03)<br>p = 0.033* | -0.04 (SE = 0.03)<br>p = 0.099 | -0.03 (SE = 0.03)<br>p = 0.288 | -0.01 (SE = 0.04)<br>p = 0.867 |

#### Verbal episodic memory

We repeated the linear mixed effects models using CVLT immediate recall score and CVLT free recall long delay score as the outcome rather than the memory z-score.

**Table S11:** Estimated coefficients along with their standard errors (SE) and p-values for the tau PET SUVR (i.e., cross-sectional effect) and the tau PET SUVR by time interaction (i.e., longitudinal effect) terms in linear mixed effects models where longitudinal CVLT immediate recall is the outcome. Each column corresponds to a separate model. * p < 0.05, ** p < 0.01.

|  | Entorhinal | Hippocampus | Parahippocampal | ITG |
| --- | --- | --- | --- | --- |
| Tau PET SUVR | -0.23 (SE = 0.14)<br>p = 0.090 | -0.26 (SE = 0.12)<br>p = 0.030* | -0.37 (SE = 0.12)<br>p = 0.002** | -0.25 (SE = 0.14)<br>p = 0.078 |
| Tau PET SUVR × Amyloid group | -0.40 (SE = 0.27)<br>p = 0.133 | -0.37 (SE = 0.22)<br>p = 0.099 | -0.55 (SE = 0.23)<br>p = 0.019* | -0.05 (SE = 0.26)<br>p = 0.852 |
| Tau PET SUVR × Time | -0.00 (SE = 0.02)<br>p = 0.865 | -0.02 (SE = 0.02)<br>p = 0.127 | -0.00 (SE = 0.02)<br>p = 0.842 | -0.02 (SE = 0.02)<br>p = 0.373 |

**Table S12:** Estimated coefficients along with their standard errors (SE) and p-values for the tau PET SUVR (i.e., cross-sectional effect) and the tau PET SUVR by time interaction (i.e., longitudinal effect) terms in linear mixed effects models where longitudinal CVLT free recall long delay is the outcome. Each column corresponds to a separate model. * p < 0.05, ** p < 0.01.

|  | Entorhinal | Hippocampus | Parahippocampal | ITG |
| --- | --- | --- | --- | --- |
| Tau PET SUVR | -0.26 (SE = 0.13)<br>p = 0.048* | -0.28 (SE = 0.11)<br>p = 0.015* | -0.35 (SE = 0.11)<br>p = 0.003** | -0.10 (SE = 0.14)<br>p = 0.467 |
| Tau PET SUVR × Amyloid group | -0.65 (SE = 0.25)<br>p = 0.011* | -0.44 (SE = 0.21)<br>p = 0.039* | -0.67 (SE = 0.22)<br>p = 0.003** | -0.15 (SE = 0.26)<br>p = 0.560 |
| Tau PET SUVR × Time | -0.00 (SE = 0.02)<br>p = 0.811 | -0.03 (SE = 0.01)<br>p = 0.028* | -0.01 (SE = 0.02)<br>p = 0.708 | 0.01 (SE = 0.02)<br>p = 0.486 |

#### Continuous amyloid burden

We repeated the linear mixed effects models using a continuous amyloid burden index, mean cortical PiB DVR, instead of the binary amyloid status. Our findings regarding the cross-sectional and longitudinal main effects of tau remained unchanged, with the exception that the association of entorhinal tau with the rate of change in CFQ became trend-level (*p* = 0.055).

**Table S13:**
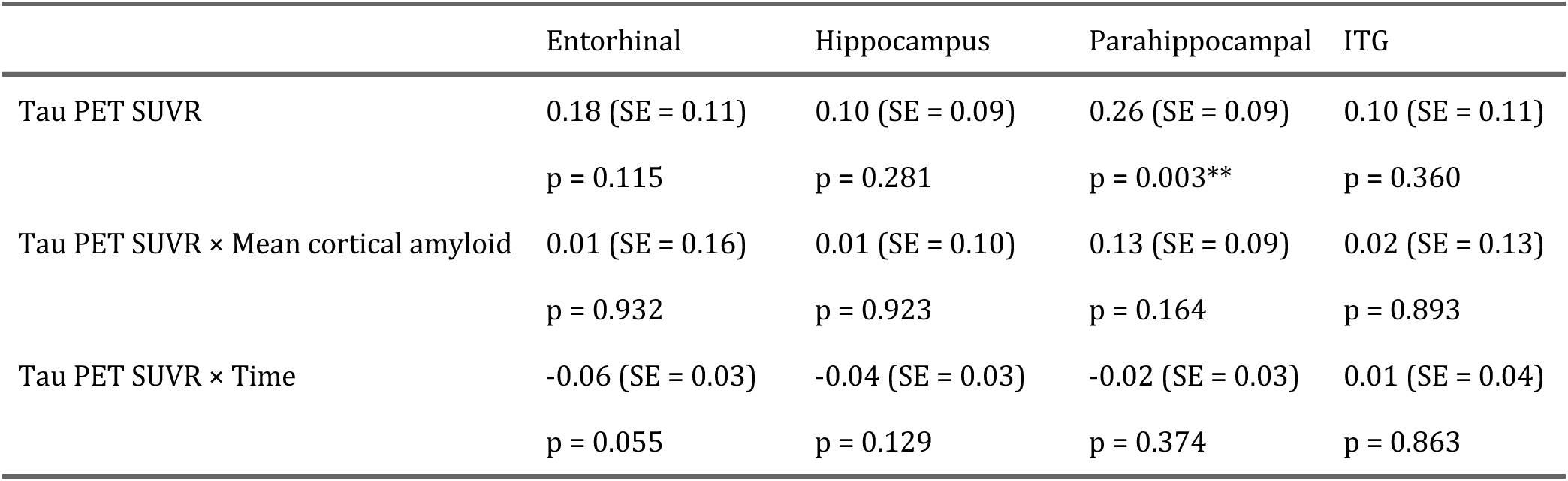
Estimated coefficients along with their standard errors (SE) and p-values for the tau PET SUVR (i.e., cross-sectional effect) and the tau PET SUVR by time interaction (i.e., longitudinal effect) terms in linear mixed effects models where longitudinal CFQ is the outcome. Instead of binary amyloid status, a continuous amyloid burden index is included as a covariate. Each column corresponds to a separate model. ** p < 0.01.

**Table S14:** Estimated coefficients along with their standard errors (SE) and p-values for the tau PET SUVR (i.e., cross-sectional effect) and the tau PET SUVR by time interaction (i.e., longitudinal effect) terms in linear mixed effects models where longitudinal memory z-score is the outcome. Instead of binary amyloid status, a continuous amyloid burden index is included as a covariate. Each column corresponds to a separate model. * p < 0.05.

|  | Entorhinal | Hippocampus | Parahippocampal | ITG |
| --- | --- | --- | --- | --- |
| Tau PET SUVR | -0.17 (SE = 0.13)<br>p = 0.188 | -0.21 (SE = 0.10)<br>p = 0.051 | -0.25 (SE = 0.10)<br>p = 0.015* | -0.19 (SE = 0.13)<br>p = 0.146 |
| Tau PET SUVR × Mean cortical amyloid | -0.20 (SE = 0.19)<br>p = 0.278 | -0.14 (SE = 0.12)<br>p = 0.237 | -0.22 (SE = 0.11)<br>p = 0.054 | -0.10 (SE = 0.14)<br>p = 0.468 |
| Tau PET SUVR × Time | -0.00 (SE = 0.02)<br>p = 0.830 | -0.03 (SE = 0.01)<br>p = 0.038* | -0.00 (SE = 0.01)<br>p = 0.760 | -0.00 (SE = 0.02)<br>p = 0.915 |

### R packages used for generating figures and tables

We used ggeffects, ggplot2, ggpubr, and GGally [50–53] to generate the figures, and gtsummary, modelsummary, and flextable [54–56] to generate the tables.

## Notes

### Competing Interest Statement

The authors have declared no competing interest.

### Author Declarations

Research protocols were conducted in accordance with United States federal policy for the protection of human research subjects contained in Title 45 Part 46 of the Code of Federal Regulations (45 CFR 46), approved by local institutional review boards (IRB) and the National Institutes of Health, and all participants gave written informed consent at each visit.

### Summary of Updates

Revised for Alz & Dem: DADM Journal submission.

